# Generating Advancements in Longitudinal Analysis in X and Y Variations: Rationale, Design, and Methods for the GALAXY Registry

**DOI:** 10.1101/2024.08.14.24311888

**Authors:** Alexandra Carl, Samantha Bothwell, Karli Swenson, Ryan Bregante, Lilian Cohen, Virginia Cover, Anna Dawczyk, Gail Decker, Stephen B. Gerken, David Hong, Susan Howell, Armin Raznahan, Alan D. Rogol, Nicole Tartaglia, Shanlee Davis

## Abstract

Sex chromosome aneuploidies (SCAs) are a family of genetic disorders that result from an atypical number of X and/or Y chromosomes. SCAs are the most common chromosomal abnormality, affecting ∼1/400 live births, yet are often underdiagnosed, leading to over-representation of more severely impacted individuals in many clinical studies. In addition to this ascertainment bias, existing work in SCAs has also been limited by low geographic and demographic diversity. To address these limitations, we have created the Generating Advancements with Longitudinal Analysis in X and Y variations (GALAXY) Registry. To date, GALAXY has accrued 295 verified SCA participants. Next steps include targeted recruitment of minoritized individuals regarding race, ethnicity, and socioeconomic status, and continuing engagement with the SCA community.

## 1. Introduction

Sex chromosome aneuploidy (SCA) conditions refer to a family of genetic disorders resulting from an atypical number of X or Y chromosomes. Together, SCAs occur in 1 in ∼400 live births, making them the most commonly occurring chromosomal abnormalities despite being under-recognized and underdiagnosed.(Nielsen & Wohlert, 1990) Supernumerary SCAs include Klinefelter syndrome (47,XXY; 1/600 males), Trisomy X syndrome (47,XXX; 1/1000 females), Jacob’s syndrome (47,XYY; 1/1000 males)^1^, and rarer tetrasomy and pentasomy conditions (e.g., 48,XXYY, 48,XXXY, 48,XXXX, 49,XXXXY, etc. estimated to occur between 1 in 17,000 to 100,000 live births)(Linden et al., 1995). Historically, SCAs have been under-ascertained with just 10-40% of individuals ever receiving a diagnosis, of whom only 10% are diagnosed in childhood(Abramsky & Chapple; Sánchez et al., 2023; Tartaglia et al., 2020).Additionally, the diagnostic odyssey often starts with an observed medical problem leading to genetic testing, which introduces ascertainment bias such that individuals with more severe presentations are more likely to be diagnosed. However, the rate of prenatal identification is increasing as prenatal cell-free DNA becomes part of routine prenatal care(Abramsky & Chapple; “Screening for Fetal Chromosomal Abnormalities: ACOG Practice Bulletin, Number 226,” 2020). With this increase in both earlier diagnosis and overall ascertainment of SCAs, there is an unprecedented demand for patient-centered research(Velvin et al., 2022) targeted at improving health and wellbeing for individuals affected by these conditions.

Several challenges hinder clinical research in SCAs. Foremost, SCAs are each rare leading to an overrepresentation of underpowered studies on small cohorts at single institutions. Due to small numbers, SCA conditions are often pooled together for more analytical power thus minimizing unique differences driven by karyotype or other genetic and environmental covariates that contribute to the phenotypic heterogeneity. While larger-scale retrospective studies have been possible using European registries(Boyd et al., 2011), these are predominately studies of adults and lack racial, ethnic, and/or socioeconomic diversity that truly represents all persons living with SCA. Newborn screening studies from the 1960s-1970s prospectively followed 304 individuals with SCAs diagnosed incidentally at birth across cohorts in Colorado(Robinson et al., 1979), Denmark(Neilson et al., 1979), Toronto(Stewart et al., 1979), New Haven(Leonard et al., 1979), Tokyo(Higurashi et al., 1979), and Edinburgh(Ratcliffe et al., 1979). Although these studies provide a foundation for the natural history of growth and development in these conditions, they fail to reflect current clinical practice and healthcare advancements over the past half-century.

To address some of these limitations, a clinical registry and robust infrastructure was developed – based on the successful Inspiring New Science in Guiding Healthcare in Turner Syndrome (INSIGHTS) Registry(Shankar et al., In Press) – to capture both clinically validated and self-reported data on individuals with supernumerary SCAs(Shankar et al., In Press). With support from patient advocacy organizations, including the Association for X and Y Variations (AXYS)(AXYS, 2023a), Living with XXY(XXY, 2019), and The XXYY Project(AXYS, 2023b), the <u>G</u>enerating <u>A</u>dvancements in <u>L</u>ongitudinal <u>A</u>nalysis in <u>X</u> and <u>Y</u> Variations (GALAXY) Registry launched in 2022. This registry is guided by a diverse Steering Committee (SC) of researchers, clinicians, self-advocates, and parents of affected individuals, to inform the development, oversight, and direction of all GALAXY projects. Through prioritizing sustainability, transparency, and minimizing participant burden, the overarching goal of the GALAXY Registry is to improve health outcomes for individuals with SCAs by serving as an infrastructure for future SCA research based on a large, heterogeneous, and longitudinal sample.

## 2. Methods

### 2.1 Structure of GALAXY

An overview of the GALAXY infrastructure is depicted in Figure 1. A Steering Committee (SC) composed of 13 collaborators – including clinicians, researchers, and family and self-advocates – was established to direct the GALAXY Registry and ensure that resource and data use reflect the interests and priorities of the SCA community. The role of the SC is to provide stakeholder input, consensus, and oversight for research projects conducted by approved study teams. The responsibilities of the SC include review, modification, and ultimate approval of study-related documents including, but not limited to, the study protocol, data collection instruments, and participant-facing materials (i.e., recruitment materials). SC members are all volunteers, who serve a two-year term, meet monthly to establish goals and priorities, review new proposals, and oversee project progress. Members follow standard conflict of interest (COI) guidelines and are cleared of COI prior to approval of membership to the SC. In addition, smaller working groups are developed as needed to accomplish specific tasks, including a Diversity, Equity, and Inclusion (DEI) group, a Genetic/Medical group, and a Psych-Behavioral Health group.

**Figure 1.**
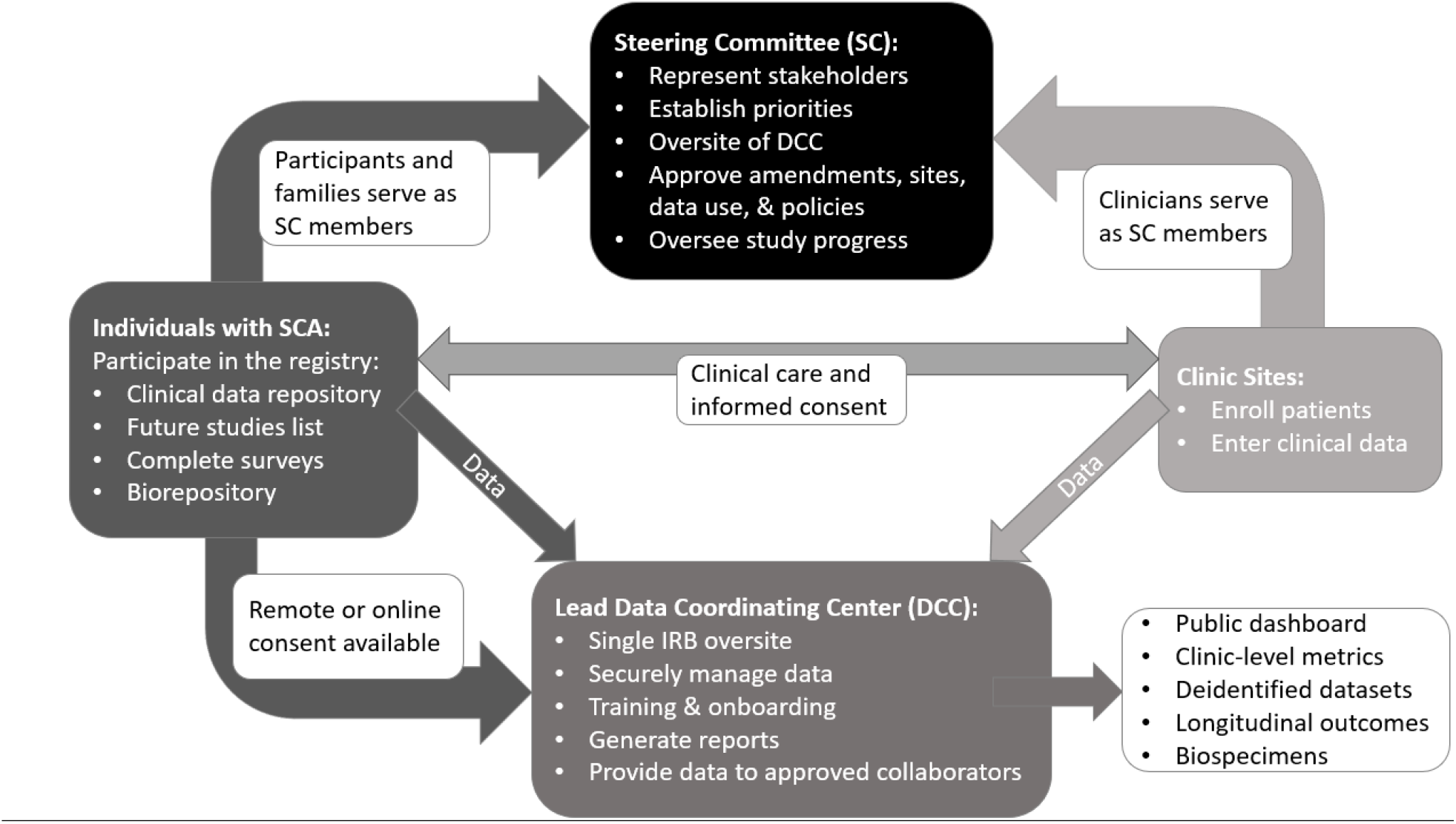
Stakeholders involved in the GALAXY registry. Abbreviations: SC=Steering Committee; SCA=sex chromosome aneuploidy; DCC=data coordinating center; IRB=institutional review board

The University of Colorado (CU) serves as the lead site and data coordinating center for the GALAXY Registry. CU is responsible for administration tasks related to the registry including regulatory compliance through a Single Institutional Review Board (sIRB), development and maintenance of the database and Standard Operating Procedures, coordinating virtual SC meetings, onboarding new participant enrollment sites, and training of all study personnel. A timeline of study start up can be seen in figure 2. REDCap (Research Electronic Data Capture)(Harris et al., 2009) is used for both clinically verified data capture and participant-facing surveys in GALAXY. REDCap is a secure, HIPAA-compliant, web-based application designed to support data capture for research studies, with a built-in e-consent framework and dual language management system to allow participants to consent in their preferred language (English or Spanish). The GALAXY Registry is approved by the Colorado Multiple Institutional Review Board (COMIRB, #20-0482).

**Figure 2.**
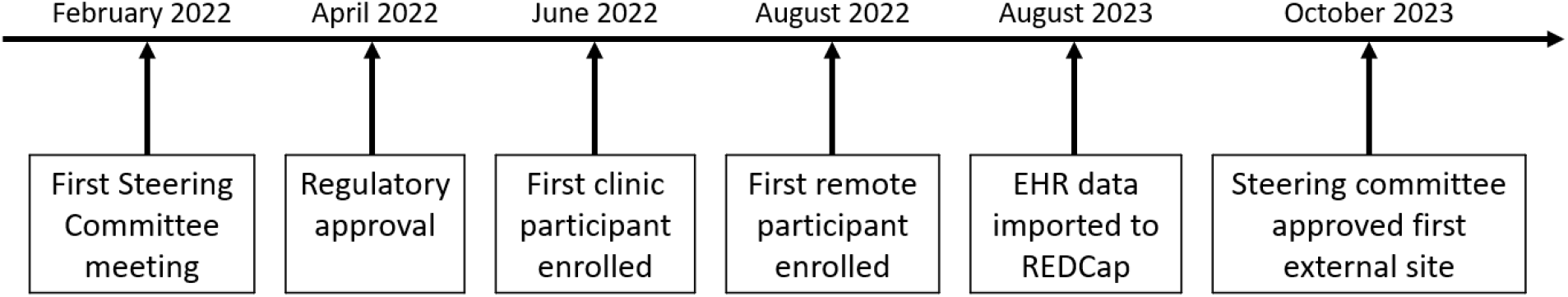
Timeline of study-start up for GALAXY. EHR=electronic health record

Individuals with SCAs can enroll into GALAXY through a participating clinic site or online with appropriate genetic test results confirming an SCA. Participating sites are approved, multidisciplinary clinics that provide care to individuals with SCAs, agree to enroll participants into GALAXY, and have resources to contribute clinical data into the database. Applications require approval by the Steering Committee to ensure the site has capacity to meaningfully contribute to the registry goals. The onboarding process includes the local IRB ceding to COMIRB, finalizing the Registry Agreement that authorizes data and material transfer between institutions, and training new site personnel.

### 2.2 Recruitment and Enrollment

At participating clinics, potentially eligible patients are identified by clinic providers, via review of past and upcoming clinic schedules by research staff, and/or via electronic health record (EHR) tools.

Inclusion criteria include:

1. Cytogenetically confirmed diagnosis of SCA;
2. Any age, gender, or language; and
3. Providing informed consent for individuals ≥18 years of age, or parent/guardian consent for individuals <18 or ≥18 with impaired decision making (plus assent for minors).

The only exclusion criterion is lack of documentation of genetic testing confirming an SCA diagnosis or deceased individuals.

Enrollment processes are outlined in Figure 3. Eligible patients and/or their legal guardians are recruited by a member of their clinic site or research team, and informed consent and assent documents are available in both English and Spanish, with the opportunity to translate into other languages as a need arises. The SC intends that participating staff approach >80% of eligible clinic patients about the study opportunity and obtain >50% participation agreement.

**Figure 3.**
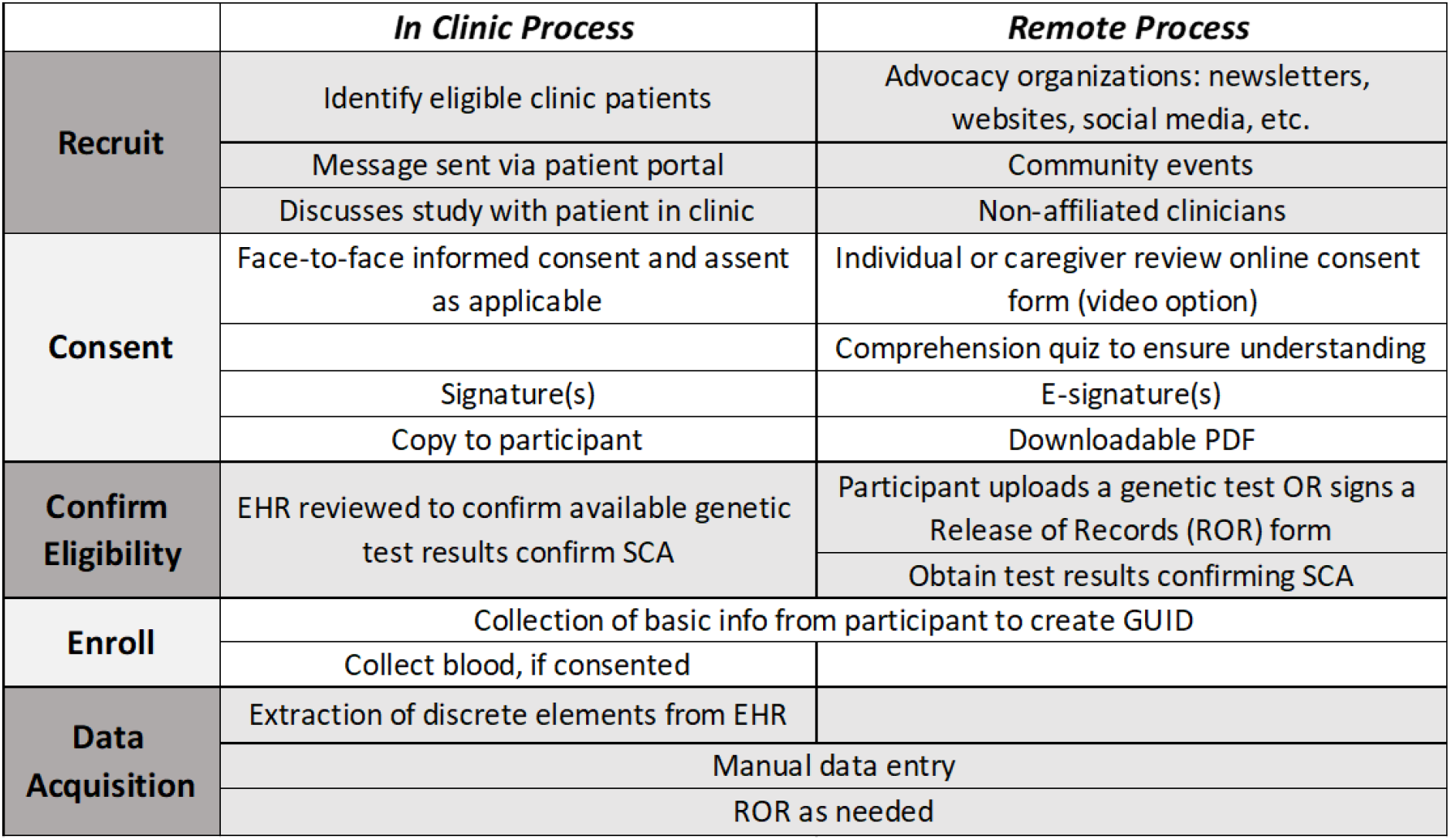
Recruitment and data acquisition processes. ROR=Release of Records: GUID=Global Unique Identifier: EHR=Electronic Health Record

Informed consent is obtained from participants 18 years and older who can consent for themselves. Informed consent is obtained from parents or legally authorized representatives of individuals under 18 years of age or those 18 years and older who are unable to consent for themselves due to developmental disability (as determined by the patient’s clinician and/or previous legal determinations in accordance with the applicable law). Additionally, assent is obtained from those between 7 and 17 years and those 18 years and older who are unable to consent for themselves.

Eligible individuals also have the option to self-enroll if they are not seen in one of the participating sites. The GALAXY Registry is advertised on institution and organization websites and social media pages. Partnerships with SCA advocacy groups such as the Association of X and Y Variations (AXYS), The XXYY Project, and Living with XXY allow broader recruitment. Research staff attend and present the GALAXY registry during virtual community meetings and webinars to talk about the registry and inform potential participants of the study procedures. Additionally, SCA advocacy groups post online about the registry on their social media platforms (e.g., Facebook groups, Instagram) and on email newsletters. In July 2023, video consenting was approved for the registry due to the observed increased incidence of language-based difficulties in SCA populations(Lee et al., 2011). Videos are available alongside written online consent that break down and explain each section of the consent for individuals for whom reading may be a barrier.

To confirm eligibility, individuals enrolling online, and not seen within a participating clinic, are required to upload their genetic test results or sign a Release of Records for the research team to request a valid copy of their cytogenetic lab report. Participants (or their legal guardian) can securely upload additional applicable medical records or provide permission for records to be requested from clinics and released to the study team so that clinically verified data can be included in the registry database. Expanding recruitment to individuals who are not patients at a participating clinic site widens participation opportunities as well as recruits a more diverse sample to the registry. The study is working to obtain General Data Protection Regulation (GDPR) compliance, which would allow individuals from the European Union to also consent through the online process. In addition to allowing medical records to be used for research purposes, participants can consent to optional procedures, including being recontacted for future SCA-related research and receiving electronic research surveys. Individuals seen at a participating clinic can additionally consent to contributing a blood sample to the biorepository.

### 2.3 Data acquisition, management, and sharing

At the time of enrollment, participants provide demographic information, including race/ethnicity, insurance status, birth date, location of birth, and current residence. Birth, medical, and surgical history, diagnostic information, and medications are abstracted directly from medical records upon enrollment and updated annually. For participants who receive care from a participating institution, discrete data elements are automatically extracted monthly from the EHR into the research database. This EHR extraction script was developed and designed by the lead site to allow standardized implementation at other participating clinic sites. Data that cannot be automatically exported are manually reviewed and entered into REDCap by the local study team. For health records outside the participating sites, a signed Release of Records is obtained during the consent process, and relevant medical records are requested, abstracted, and entered into the database manually. When data cannot be obtained from medical records, the study team contacts the participant for additional information. Data quality is ensured by trained research staff following Standard Operating Procedures and data are validated during site onboarding and training.

Data sharing from the GALAXY Registry is managed following the Data Sharing Agreements and Data Use Policy approved by the Steering Committee and publicly available (https://redcap.ucdenver.edu/surveys/?dashboard=YHT3DR8JYX4). In addition to requesting data, investigators can apply to disseminate recruitment materials to participants who consented into the optional Future Studies Recruitment List (Figure 4). The Steering Committee reviews all proposals to ensure they are of sound scientific merit and beneficial to individuals and families in the SCA community. Data proposals are requested to include community engagement and describe planned dissemination to the SCA community, separate from academic conferences and peer-reviewed publication.

**Figure 4.**
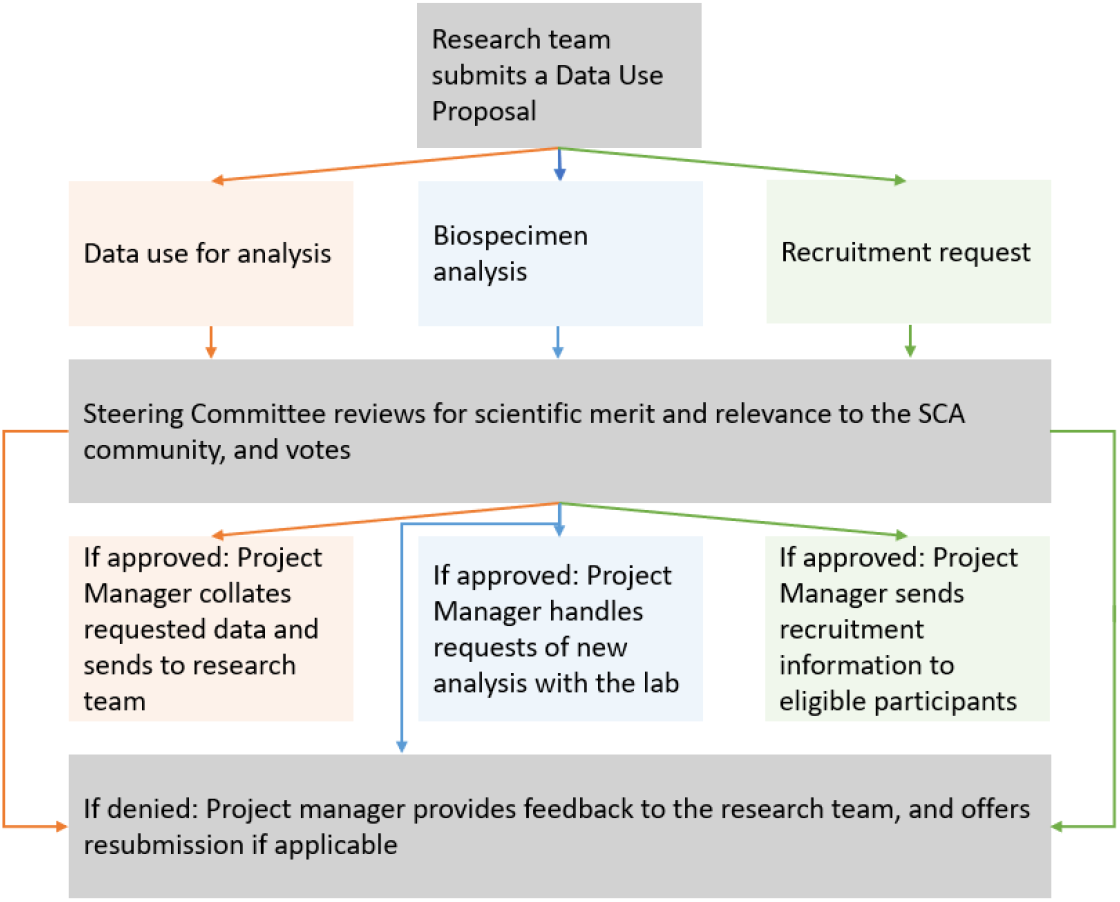
How reieurcherj cun inter jet with GALAXY

In addition, each participant is given a Global Unique Identifier (GUID) through the National Institutes of Health (NIH) Biomedical Research Informatics Computing System. The Rare Diseases Registry Program (RaDaR) provides guidance for setting up and maintaining registries for rare diseases. The GUID and common data elements taken from RaDaR facilitate data sharing without sharing personally identifying information (PII) across studies.

Sharing data outside the academic community is a goal of the GALAXY registry. A live dashboard with basic data is publicly available to increase transparency with the registry participants, prospective researchers, and the SCA community. Additionally, presentations and publications about the registry are shared on the study website (galaxyregistry.org). The consent form is also publicly available (redcap.link/GALAXY-consent) for individuals and prospective researchers to review.

### 2.4 Outcome measures

Common data elements (CDEs) were selected from NIH repositories. Expanded and specialty-specific data elements were developed and refined by interdisciplinary members of the SC to ensure reliability and validity of the measures. Validated patient-facing measures are included (supplemental material) and any non-validated questionnaires were reviewed and modified by the SC and pilot tested with individuals with SCAs.

Data elements in GALAXY, taken from participant’s EHR:

1. Genotype
2. Timing and reason for SCA diagnosis
3. Other medical diagnoses (i.e., presence, number, and type)
4. Growth parameters and vital signs
5. Documented physical exam including Tanner staging, dysmorphic features, and neurological exam
6. Birth and perinatal history
7. Surgical procedures (i.e., presence, number, and type)
8. Hospitalizations (i.e., presence, number, reason)
9. Treatments including medications (i.e., presence, number, and type)
10. Imaging tests performed and results
11. Laboratory tests performed and results
12. Psychological and cognitive tests performed and results Additionally, from those who consent to participate in the optional questionnaires and biorepository:
13. Patient-reported outcomes from questionnaires in domains of quality of life, wellbeing, lived experiences, sociodemographic information, access to care, psychosocial health, etc. (Supplemental Table 1)
14. Blood draw with plasma, serum, and buffy coat stored in the biorepository

GALAXY surveys and information regarding future research opportunities are sent via REDCap to the email address the participant provided at enrollment. For participants under 18 years--or at the request of an adult participant--the emails are sent to a parent/guardian. Surveys are electronically administered utilizing the survey queue tool in REDCap and questionnaires specific to the participant’s age or karyotype are populated into their unique survey queue, which participants can return to at any time.

Surveys are designed to collect participant-reported outcomes including sociodemographic information, access and experience with clinical care, quality of life, wellbeing, and psychosocial health. Surveys include a statement indicating results are not shared with the participant’s clinical providers, and that participants should contact their doctor(s) directly if they have clinical concerns that arise from completing the survey.

### 2.5 Statistical Analysis

Demographic summaries are presented as count (%) and stratified by type of enrollment (i.e., in-person at Children’s Hospital Colorado vs online). Data visualization of the density of patient enrollment by state was generated in R, version 4.4.0, and is similarly stratified by type of enrollment.

## 3. Results

Children’s Hospital Colorado (CHCO) began recruiting clinical patients in June of 2022; four more sites are currently onboarding as of July 2024. A total of 295 individuals have been enrolled and validated based on enrollment criteria into the registry as of June 25, 2024, 188 from CHCO and 107 from self-enrollment with an average of 14 new participants per month. The median age at enrollment is 10.7 years (IQR : 2 – 17.9 years), ranging from 0 to 87 years. Demographic data can be seen in Table 1. Forty-one total states are represented as visualized in Figure 5, as the state of residence for US-based participants.

**Table 1.**
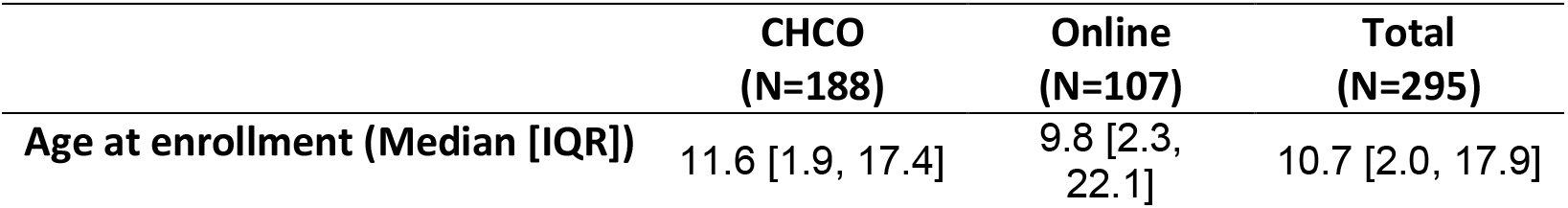
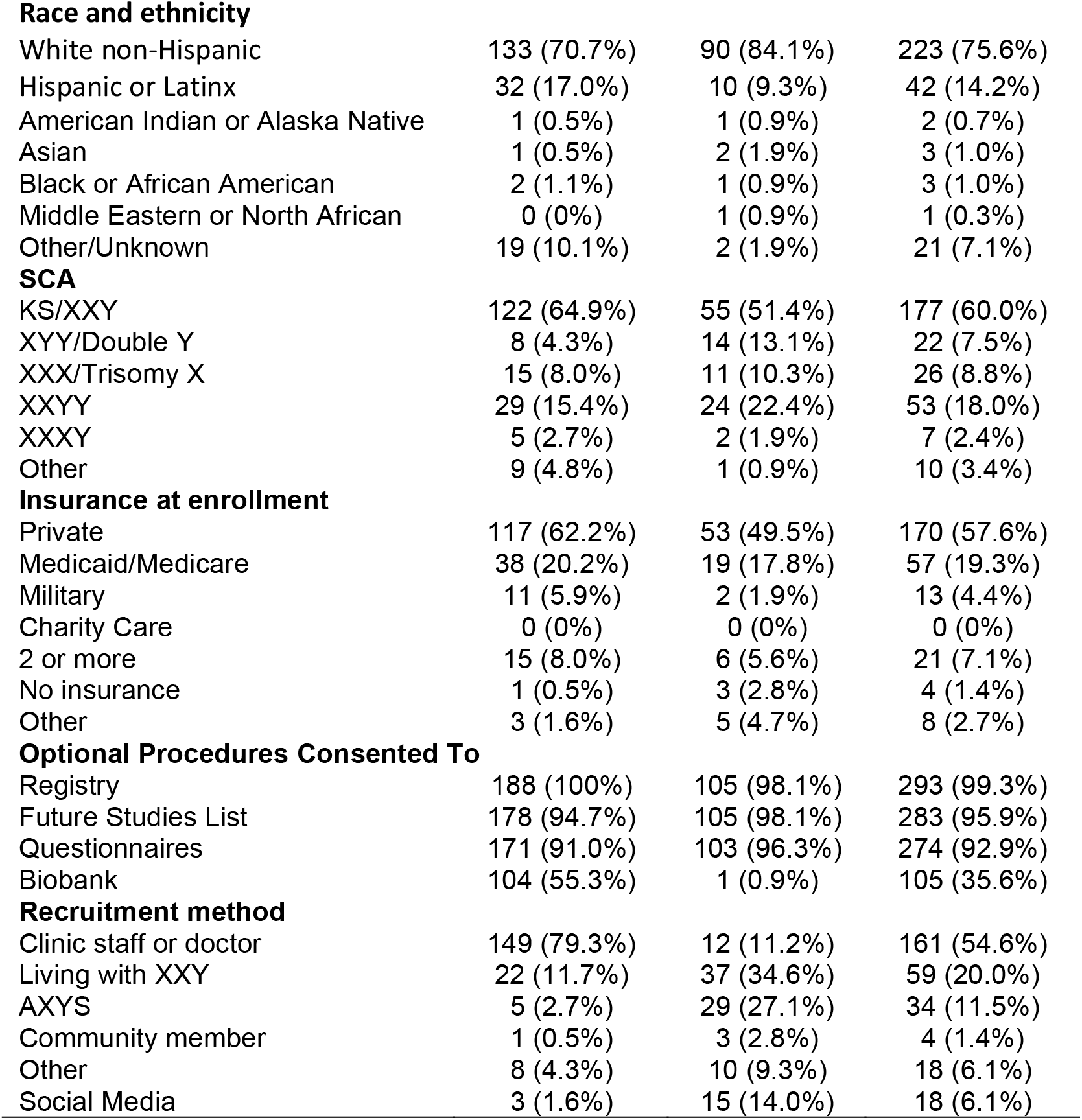
Sample demographic characteristics.

**Figure 5.**
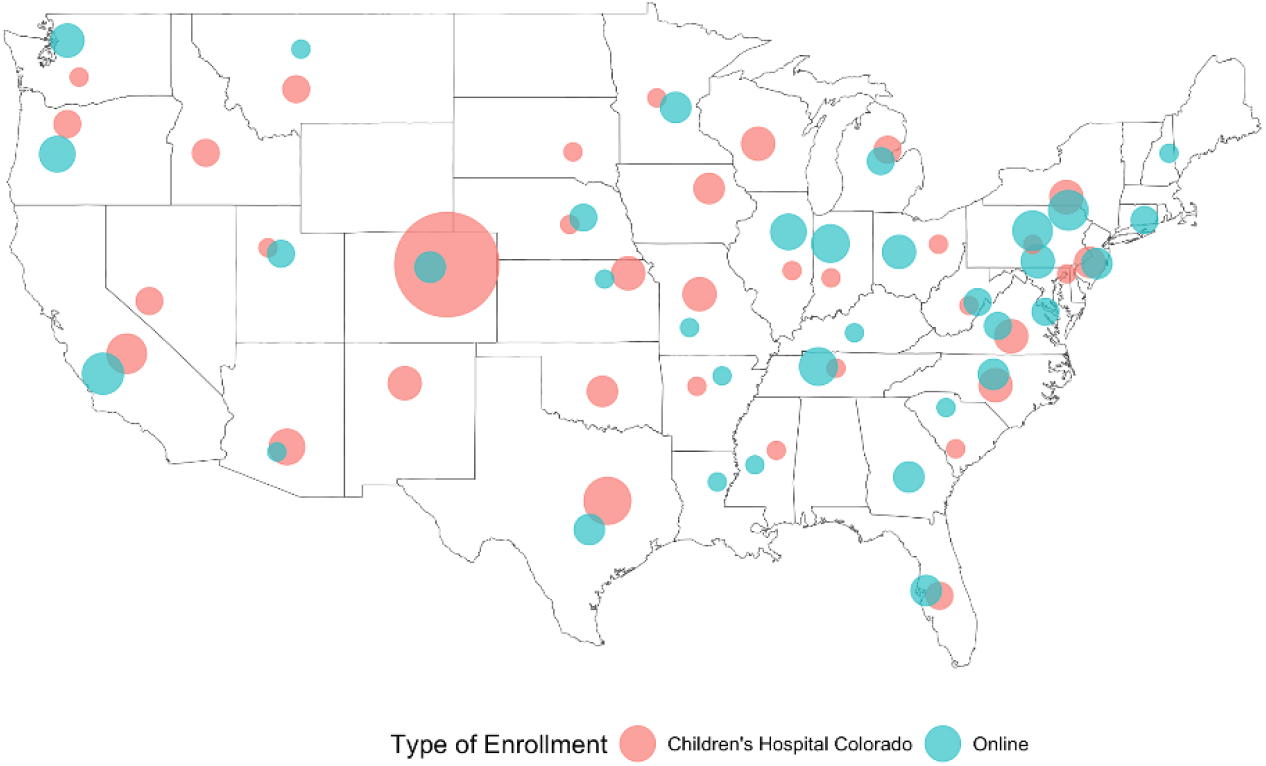
Density of participants enrolled in each state by type of enrollment; larger circles represent larger patient populations. Maximum recruitment for a single state is 99 patients in Colorado. Points are jittered within a state and do not reflect recruitment from individual cities.

## 4. Discussion

The GALAXY Registry is a collaborative effort among invested stakeholders to collect and store longitudinal clinical and self-reported data from individuals with SCA in a centralized international database with common, expanded, and specialized data elements. Since launching in April 2022, GALAXY has enrolled >250 individuals from diverse genetic, geographical, racial, and socioeconomic populations and continues to enroll participants and collect data, with the expectation of serving as a sustainable resource for future natural history and other observational SCA studies.

Rare disease registries are recognized as important contributors to evidence-based medicine and are important for describing natural history and heterogeneity of rare conditions to informing clinical care guidelines(Lee et al., 2011). Registries also serve as a valuable recruitment source for observational and interventional studies. Prior natural history cohorts in the SCA population have been limited to single or a few institutions and comprised of motivated participants willing and able to travel for participation in research, as well as those who historically received a diagnosis(Gutin et al., 2012). The GALAXY Registry is novel in that it is driven by a team of stakeholders from the SCA community, including patients, families, and clinicians, and enables participants to enroll in their local clinics or online. Data obtained directly from the EHR ensure that the database contains clinically validated data, promoting reliability of results. The goal of the registry is to provide a greater opportunity for patient-engagement with research within the SCA community and expand access to research for participants from a wider range of ages, geographic locations, and economic resources to meaningfully contribute to research without undue burden on the participants or families.

### 4.1 Limitations

The initial limitation is that enrollment is contingent on review of a genetic test result, which excludes individuals who have not received a clinical diagnosis (at least half of all individuals with an SCA)(Bojesen et al., 2003) and also presents a barrier for those with a diagnosis who do not have access to prior results. The current sample is heavily weighted toward younger participants with 47,XXY, which minimizes the utility of the current dataset to answer questions about aging or inform rarer SCA conditions. However, we anticipate the absolute numbers of other SCA conditions and older participants will increase over time.

The EHR, even when verified by physicians, may still contain errors of omission or commission that may impact the quality of the data. Similarly, not all variables of interest are clinically obtained, and clinical records are not always available. Despite these limitations, GALAXY represents an important resource for future SCA research as it is the first and largest clinically validated registry for this population.

### 4.2 Future directions

GALAXY will expand to additional clinical sites; four are currently onboarding and more have expressed interest. Increased national clinic representation in the registry may aid in diversifying the participant sample, including more non-XXY individuals and greater age and racial diversity. The Association for X and Y Variations (AXYS) has brought together the AXYS Clinic and Research Consortium (ACRC), which is a dedicated group of international SCA clinics. GALAXY is committed to recruiting dedicated clinic sites through this group to expand the registry’s potential and increase diversity of participants. The CHCO sample is more diverse than our online population, indicating that online recruitment is likely reaching individuals with easier access to medical research. Increasing the number of clinical sites recruiting may assist with increased diversity, and more online methods targeting historically underrepresented groups will be undertaken such as partnering with community efforts, education about research, and increased transparency.

Additionally, collaboration with learning health systems, such as PEDSnet, may facilitate collection of additional discrete data elements. A lack of funding for sites to enter data, such that data collection is dependent on investigator time, is a barrier to complete data. Additional sources of funding continue to be explored to support data entry and expansion of the registry. Ideally, funding would allow expansion to include confirmatory genetic testing for individuals who do not have results available to remove this barrier to enrollment. As the GALAXY Registry grows, we anticipate that it will serve as a powerful resource for statistical analyses aimed at clarifying the natural history of SCA and defining predictors of individual-level outcomes.

## 5. Conclusions

The GALAXY Registry – a clinical research registry for individuals with SCAs – was developed with multiple stakeholder perspectives and is diverse and growing. GALAXY provides opportunities for the SCA community to participate in clinical research regardless of their geographic location or ability to access a specialty clinic. GALAXY is building the infrastructure to serve as a resource to connect participants and researchers in the SCA community through mutual partnership and increased opportunities for research. Ultimately, GALAXY aims to provide a conduit for future longitudinal natural history research and clinical trials that will improve care and outcomes for individuals with SCAs.

## Data Availability

All data produced in the present study are available upon reasonable request to the authors

## Notes

### Competing Interest Statement

The authors have declared no competing interest.

### Funding Statement

This study did not receive any funding

### Author Declarations

Ethics committee, Colorado Multiple Institutional Review Board (COMIRB), of the University of Colorado Anschutz Medical Campus gave ethical approval for this work

## References

Abramsky, L., & Chapple, J. 47,XXY (Klinefelter syndrome) and 47,XYY: estimated rates of and indication for postnatal diagnosis with implications for prenatal counselling. Prenatal Diagnosis, 17(4), 363–368. 10.1002/(sici)1097-0223(199704)17:4<363::aid-pd79>3.0.co;2-o

AXYS. (2023a). Association of X and Y Chromosome Variations. https://genetic.org/

AXYS. (2023b). The XXYY Project and Information about 48-XXYY Syndrome. https://genetic.org/variations/about-xxyy/

Bojesen, A., Juul, S., & Gravholt, C. H. (2003). Prenatal and postnatal prevalence of Klinefelter syndrome: a national registry study. The Journal of clinical endocrinology and metabolism, 88(2), 622–626. 10.1210/jc.2002-021491

Boyd, P. A., Loane, M., Garne, E., Khoshnood, B., Dolk, H., & group, E. w. (2011). Sex chromosome trisomies in Europe: prevalence, prenatal detection and outcome of pregnancy. Eur J Hum Genet, 19(2), 231–234. 10.1038/ejhg.2010.148.

Gutin, L. S C., Bakalov, V. K., & Bondy, C. (2012). Trends in GH use in a Turner syndrome natural history study. Pediatric endocrinology reviews, 9, 725–727.

Harris, P. A., Taylor, R., Thielke, R., Payne, J., Gonzalez, N., & Conde, J. G. (2009). Research electronic data capture (REDCap)--a metadata-driven methodology and workflow process for providing translational research informatics support. J Biomed Inform, 42(2), 377–381. 10.1016/j.jbi.2008.08.010

Higurashi, M., Iijima, K., & Ikeda, U. (1979). Chromosome Survey of Newborn Infants in Tokyo: Follow-Up Study for XXY. In Sex Chromosome Aneuploidy: Prospective Studies on Children (Vol. XV, pp. 161–174). Alan R Liss Inc.

Lee, N. R., Lopez, K. C., Adeyemi, E. I., & Giedd, J. N. (2011). Chapter Six - Sex Chromosome Aneuploidies: A Window for Examining the Effects of the X and Y Chromosomes on Speech, Language, and Social Development. In D. J. Fidler (Ed.), International Review of Research in Developmental Disabilities (Vol. 40, pp. 139–180). Academic Press. 10.1016/B978-0-12-374478-4.00006-X

Leonard, M. F., Schowalter, J. E., Landy, G., Ruddle, F. H., & Lubs, H. A. (1979). Chromosomal Abnormalities in the New Haven Newborn Study: A Prospective Study of Development of Children With Sex Chromosome Anomalies. In A. Robinson, H. A. Lubs, & D. Bergsma (Eds.), Sex Chromosome Aneuploidy: Prospective Studies on Children (Vol. XV, pp. 115–160). Alan R Liss Inc.

Linden, M. G., Bender, B. G., & Robinson, A. (1995). Sex chromosome tetrasomy and pentasomy [Article]. Pediatrics, 96(4), 672+. https://link-gale-com.proxy.hsl.ucdenver.edu/apps/doc/A17519296/ITOF?u=denison&sid=bookmark-ITOF&xid=b2d038db

Neilson, J., Sillesen, I., Sorensen, A. M., & Sorensen, K. (1979). Follow-up until age 4 to 8 of 25 unselected children with sex chromosome abnormalities compaired with sibs and controls. In A. Robinson, H. A. Lubs, & D. Bergsma (Eds.), Sex Chromosome Aneuploidy: Prospective Studies on Children (Vol. XV, pp. 15–74). Alan R Liss Inc.

Nielsen, J., & Wohlert, M. (1990). Sex chromosome abnormalities found among 34,910 newborn children: results from a 13-year incidence study in Arhus, Denmark. Birth Defects Orig Artic Ser, 26(4), 209–223.

Ratcliffe, S. G., Axworthy, D., & Ginsborg, A. (1979). The Edinburgh Study of Growth and Development in Children With Sex Chromosome Abnormalities. In A. Robinson, H. A. Lubs, & D. Bergsma (Eds.), Sex Chromosome Aneuploidy: Prospective Studies on Children (Vol. XV, pp. 243–260). Alan R Liss Inc.

Robinson, A., Puck, M., Pennington, B., Borelli, J., & Hudson, M. (1979). Abnormalities of the sex chromosomes: A psrospective study on randomly identified newborns. In A. Robinson, H. A. Lubs, & D. Bergsma (Eds.), Sex Chromosome Aneuploidy: Prospective Studies on Children (Vol. XV, pp. 203–242). Alan R Liss Inc.

Sánchez, X. C., Montalbano, S., Vaez, M., Krebs, M. D., Byberg-Grauholm, J., Mortensen, P. B., Børglum, A. D., Hougaard, D. M., Nordentoft, M., Geschwind, D. H., Buil, A., Schork, A. J., Thompson, W. K., Raznahan, A., Helenius, D., Werge, T., & Ingason, A. (2023). Associations of psychiatric disorders with sex chromosome aneuploidies in the Danish iPSYCH2015 dataset: a case-cohort study. Lancet Psychiatry, 10(2), 129–138. 10.1016/s2215-0366(23)00004-4

Screening for Fetal Chromosomal Abnormalities: ACOG Practice Bulletin, Number 226. (2020). Obstet Gynecol, 136(4), e48–e69. 10.1097/aog.0000000000004084

Shankar, R. K., Carl, A. E., Law, J., Bamba, V., Brickman, W. J., Prakash, S. K., Dowlut-McElroy, T., Howell, S., Gutmark-Little, I., Klein, K., Pinnaro, C. T., Ranallo, K., Good, M., & Davis, S. M. (In Press). Inspiring New Science to Guide Healthcare in Turner Syndrome: Rationale, Design, and Methods for the InsighTS Registry. Am J Med Genet.

Stewart, D. A., Netley, C. T., Bailey, J. D., Haka-Ikse, K., Platt, J., Holland, W., & Cripps, M. (1979). Growth and Development of Children With X and Y Chromosome Aneuploidy: A Prospective Study. In A. Robinson, H. A. Lubs, & D. Bergsma (Eds.), Sex Chromosome Aneuploidy: Prospective Studies on Children (Vol. XV, pp. 75–114). Alan R Liss Inc.

Tartaglia, N., Howell, S., Davis, S., Kowal, K., Tanda, T., Brown, M., Boada, C., Alston, A., Crawford, L., Thompson, T., van Rijn, S., Wilson, R., Janusz, J., & Ross, J. (2020). Early neurodevelopmental and medical profile in children with sex chromosome trisomies: Background for the prospective eXtraordinarY babies study to identify early risk factors and targets for intervention. Am J Med Genet, 184(2), 428–443. 10.1002/ajmg.c.31807

Velvin, G., Hartman, T., & Bathen, T. (2022). Patient involvement in rare diseases research: a scoping review of the literature and mixed method evaluation of Norwegian researchers’ experiences and perceptions. Orphanet J Rare Dis, 17(1), 212. 10.1186/s13023-022-02357-y

Xxy, L. w. (2019). Living with XXY. https://livingwithxxy.org/

